# Distinguishing L and H phenotypes of COVID-19 using a single x-ray image

**DOI:** 10.1101/2020.04.27.20081984

**Authors:** Mohammad Tariqul Islam, Jason W. Fleischer

## Abstract

Recent observations have shown that there are two types of COVID-19 response: an H phenotype with high lung elastance and weight, and an L phenotype with low measures^1^. H-type patients have pneumonia-like thickening of the lungs and require ventilation to survive; L-type patients have clearer lungs that may be injured by mechanical assistance^2,3^. As treatment protocols differ between the two types, and the number of ventilators is limited, it is vital to classify patients appropriately. To date, the only way to confirm phenotypes is through high-resolution computed tomography^2^. Here, we identify L- and H-type patients from their frontal chest x-rays using feature-embedded machine learning. We then apply the categorization to multiple images from the same patient, extending it to detect and monitor disease progression and recovery. The results give an immediate criterion for coronavirus triage and provide a methodology for respiratory diseases beyond COVID-19.

---

It is commonly thought that COVID-19 presents as other coronavirus diseases, with Acute Respiratory Distress Syndrome (ARDS) caused by pneumonia-like thickening of the lungs^4^. However, recent observations suggest that half of all patients present differently, with near-normal breathing mechanics, clearer lungs, and hypoxia caused by poor oxygen transfer to the capillaries^1,2,5^. The ARDS-like H phenotype is characterized by *high* lung elastance, *high* weight, and *high* recruitment^2^ (potential to re-expand collapsed lung tissue, e.g. closed alveoli^6^); these patients require mechanical ventilation to survive^7^. The corresponding measures for the L phenotype are low^2^; applying high-pressure ventilation in these cases can injure lungs and precipitate a rapid worsening to ARDS^3,5^. It is vital to classify patients appropriately.

To date, the only way to confirm the phenotypes is through high-resolution computed tomography (CT, Fig. 1)^2^. This is inhibitory for rapid triage and impossible for hospitals with limited resources. Here, we introduce a machine-learned early detection system to identify COVID-19 and differentiate its L and H phenotypes using conventional chest x-rays.

**Figure 1.**
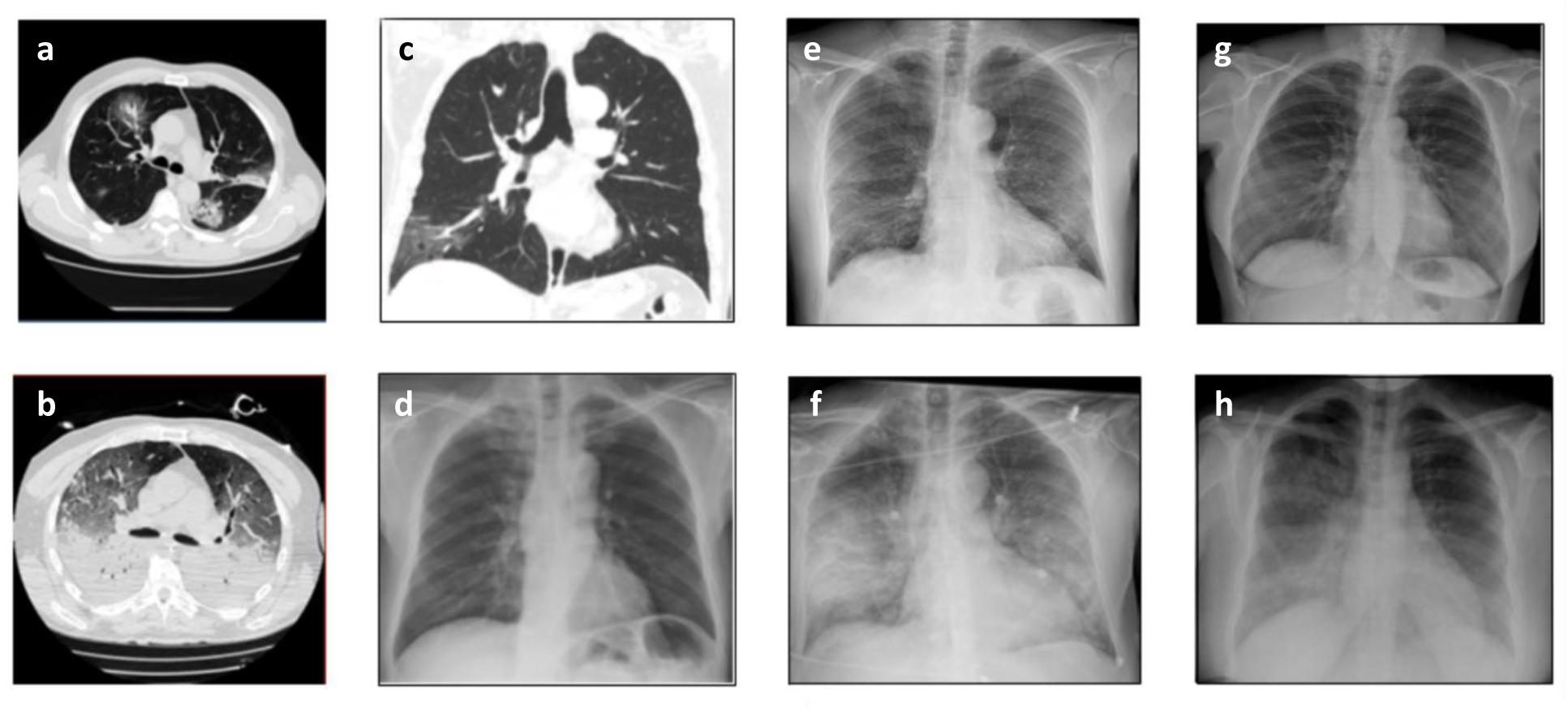
Computed tomography and x-ray data. (a,b) High-resolution CT of COVID-19 patient at (a) admission to the hospital (b) 7 days later. (c,d) Posteroanterior CT (c) and x-ray (d) images of COVID-19 patient on same day. (e,f) x-ray data of a COVID-19 patient at (e) admission, (f) 2 days later. (g) Normal x-ray. (g) x-ray of patient with viral pneumonia. (a,b) taken from [2]. (c,d) taken from [15] (e-h) taken from [12].

That this is possible comes from the origin of CT images: they are made from a collection of x-rays taken at different angles, each of which contains information on lung size, shape, and density. Resolution of CT images improves with the number of different views; higher resolution removes artifacts and provides cross-sections that allow visual separation of internal organs (Figs. 1a-c). The significance of this can be seen by comparing CT scans with individual x-ray images (Figs. 1e-h): each x-ray picture is a 2D projection of the 3D body image, so that all internal features are superimposed. This overlap complicates diagnosis considerably, especially when disease forms around the heart and behind the breastbone. Nevertheless, frontal x-rays are the most frequent radiological test performed in intensive care units^8^ and are used often at the initial point of care to diagnose respiratory distress, e.g. pneumonia and chronic obstructive pulmonary disease^9^.

The images in Figs. 1d-h, taken from the publicly available COVIDx dataset^10^, show representative chest x-rays from normal, pneumonia-positive^11^, and COVID-19-positive patients^12^. Compared with a normal image, a pneumonia x-ray shows a diffuse pattern characteristic of fluid-filled lungs. Known as ground-glass opacities when translucent and consolidation when fully opaque, these regions form when an overstimulated immune system fills the alveoli with cells and fluid^13,14^. Consequently, the location and density of these regions give many indications of the type and stage of disease. Similar patterns are seen in COVID-19 patients^15^, particularly as the illness progresses (Figs. 1b,d,f).

Figure 2 shows the results of three different numerical approaches for embedding features from the x-ray images. Each reduces the high-dimensional image space to a 2D plot of the samples. All are unsupervised, in the sense that data labels are known but only provided afterward for visualization; they are not seen or used by the algorithms. Figure 2a shows the output from Principal Component Analysis (PCA), which is a linear technique that rearranges the input data to maximize variance. It preserves large distances between points, so that samples that are distinct from each other end up far apart. It gives three clusters of x-rays with significant overlap among the points. Projection along the main principal component (Fig. 2b) gives a Gaussian profile for COVID-19 patients, a direct result of the broad statistics of the underlying data.

**Figure 2.**
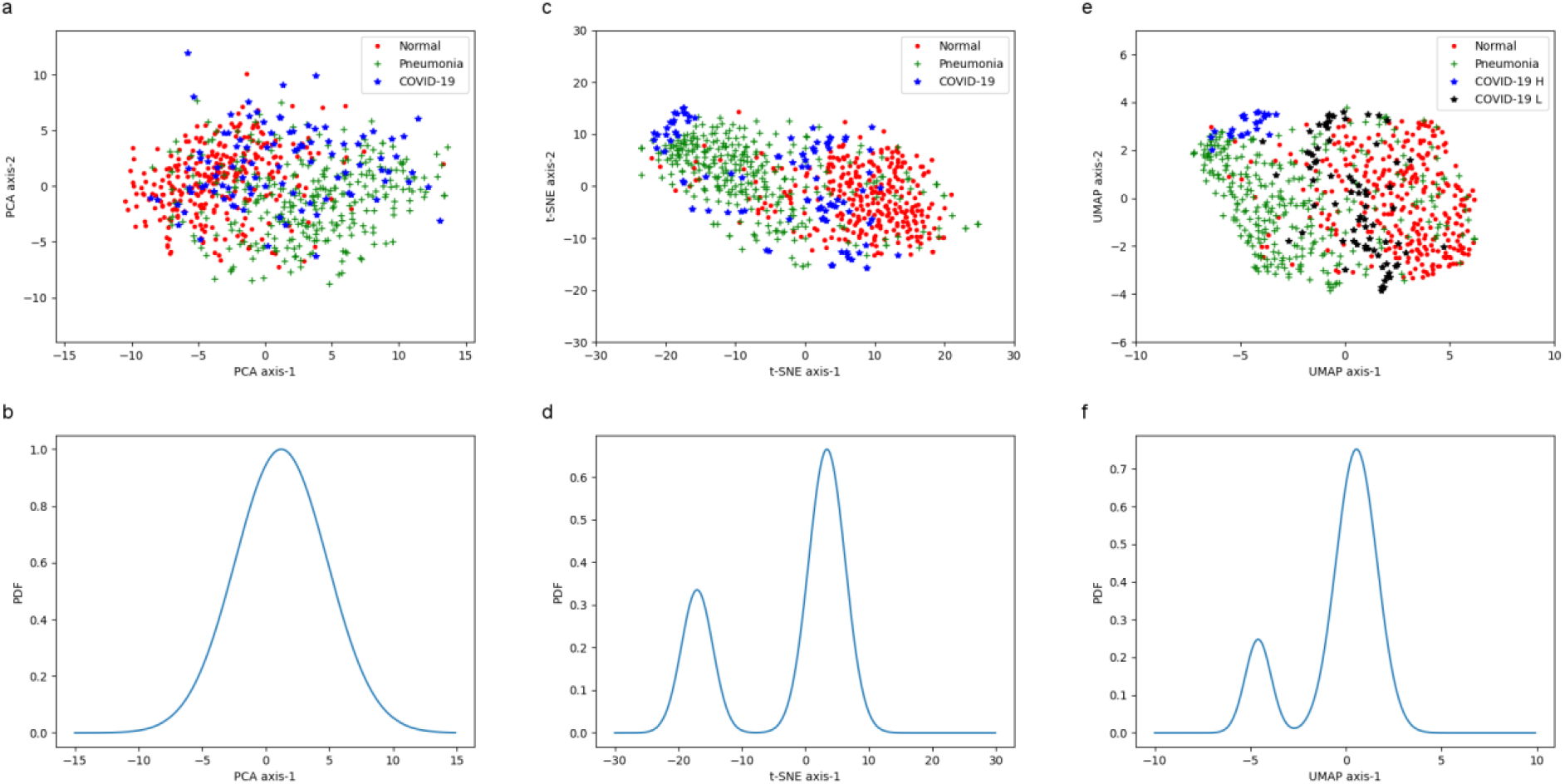
Clustering of COVID-19 patients from frontal (PA) x-ray images. Top row: Scatter plots of different algorithms. Each sample point is an x-ray image and was labeled for visualization only; labels were not used for training. Bottom row: projections. (a,b) Principal Component Analysis. (c-f) Feature-embedded machine learning. (c,d) t-SNE, (e,f) UMAP.

In contrast, Stochastic Neighbor Embedding (SNE) is a nonlinear mapping that emphasizes similarity between sample points, e.g. the joint probability of having features in common^16^. Figures 2c,d show the results of SNE using Student’s t-test as a metric (Gaussian distributions for relative probability)^17^. Remarkably, the network organizes the COVID-19 patients into two distinct clusters: one that resembles pneumonia and one that has a more normal presentation.

Unfortunately, the COVID image dataset^12^ has no labels for the L and H phenotypes. However, most chest x-rays have radiologist and physician notes on the stage of disease and response to treatment. This metadata, discussed below and in the Supplementary Information, supports the labeling of t-SNE data according to phenotype.

One problem with t-SNE clustering is that the mapping only gives information on similarity within a group, with no measure of dissimilarity between them. Neither the size of the clusters nor the distance between them has any physical meaning^18^. A more sophisticated clustering algorithm that overcomes these issues is Uniform Manifold Approximation and Projection (UMAP), which categorizes objects using Riemannian geometry and algebraic topology^19^. It is a *k*-neighbor graph-based algorithm in the same class as t-SNE and again gives two clusters (Figs. 2e,f). Compared with the t-SNE plot, the UMAP plot has tighter groupings and sharper boundaries between the x-ray types. As a consistency check, we use the L and H labels from the t-SNE clusters to color the UMAP (again, this is for visualization purposes only and was not used by the algorithm). This confirms that UMAP separates the same points as t-SNE, but with a better mapping.

A provocative feature of both mappings is the isolation of the H phenotype from the pneumonia samples, indicating a unique signature of COVID-19 infection. Similarly, the placement of L-type patients between the reference ones indicates identifiable lung damage in the x-ray image, reconciling the simultaneous observations of normal breathing and low oxygenation levels^1,5^. Future work will look for signs of susceptibility to ventilation-induced lung injury^3,20^ (a diagnostic not currently available^20^) and test the hypothesis that COVID-19 is a different class of coronavirus from those that cause MERS and SARS^21^.

As UMAP gives quantitative distance metrics, and accommodates new data better than t-SNE^19^, we focus analysis on the UMAP mapping. As a first step, we observe that the groupings in Fig. 2e are nearly orthogonal; re-plotting the data in terms of these major axes gives the representation in Fig. 3a. The two clusters of data are bound by distinct ellipses, each containing 95% of their identified x-rays. The vertical L cluster is broader and more disperse than the horizontal H cluster, with external points on both sides of its major axis. From the patient metadata^10^, it was found that the isolated point in the upper left is from the youngest patient in the dataset. The outlying point in the upper right has a partner x-ray image inside the L ellipse, taken from the same patient at an earlier time. The “misclassified” point in the H cluster also has an earlier partner inside the L ellipse. The remaining external point, bisecting the ellipses, has ambiguous radiology and no clear marker suggesting outlier status.

**Figure 3.**
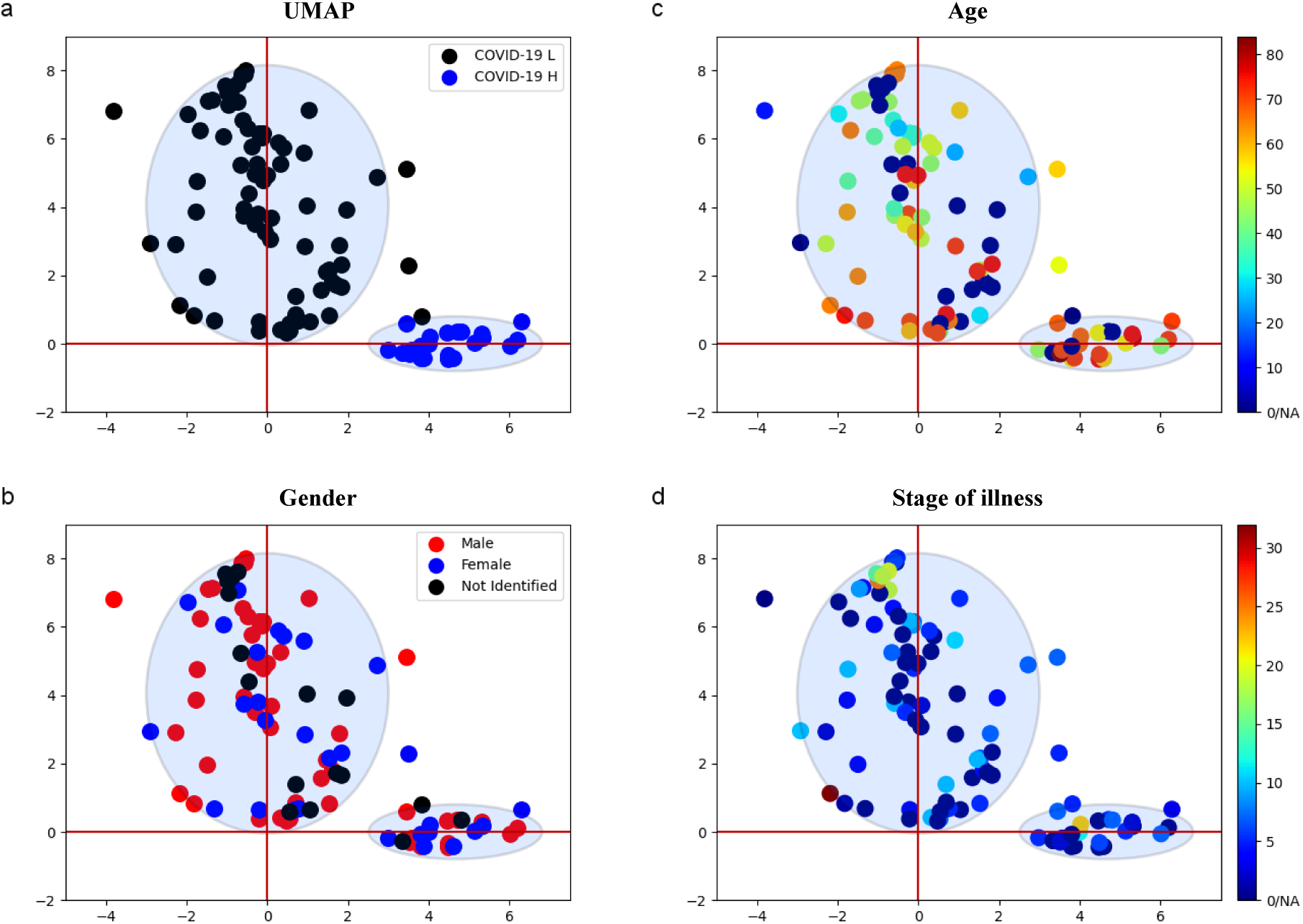
Classification of L and H phenotypes using UMAP clusters. (a) Re-mapped plot of Fig. 2e using principal axes. (b) Gender. (c) Age. (d) Stage of illness (measured in days from first hospitalization or sign of infection). Ellipse boundaries indicate 95% confidence levels.

The algorithms treat each x-ray independently and have no knowledge that multiple images could belong to the same patient. When clustering images within a set, the two methods have different priorities: t-SNE emphasizes commonality, while UMAP takes into account both locally similar features and globally different ones. This difference explains the appearance of an “L” point in the H cluster and, combined with a distance metric, suggests that UMAP can provide a scale of disease severity. While the limited data prevents its development here, patient progression and recovery are captured: worsening condition appears as movement from left to right in parameter space (normal- to pneumonia-like), while improvement goes in the opposite direction. Significantly, this ordering is preserved when UMAP is applied to new test data as well. Details are given in the Supplementary Information.

A down side of this and all machine learning techniques is that it is not clear what the machine is learning. Here, it could be the presence and density of opacities^14^, different rib thickness^22,23^ or heart size (from inflammation or strain^24^), or some other as-yet-unknown marker. In Figs. 3b-d, we label the points according to the confounding factors of gender, age, and stage of infection (measured in days from first hospitalization or sign of illness). The corresponding histograms are shown in Fig. 4. There are more males than females identified in the COVID-19 data, but they are dispersed in equal measure throughout the clusters (Figs. 3b, 4a,b). Labeled H-type patients are on average 20 years older than L-type patients (Figs. 3c, 4c,d), but their x-rays were taken at similar stages of development (Figs. 3d, 4e,f). Interestingly, both groups were intubated at the same rate (Figs. 4g,h), but the survival rate of L-type patients is higher (Figs. 4i,j). These distributions are consistent with observations of disease severity^25^ and re-affirm the distinct epidemiology of COVID-19 from other viral pandemics, such as the 1918 flu^26^.

**Figure 4.**
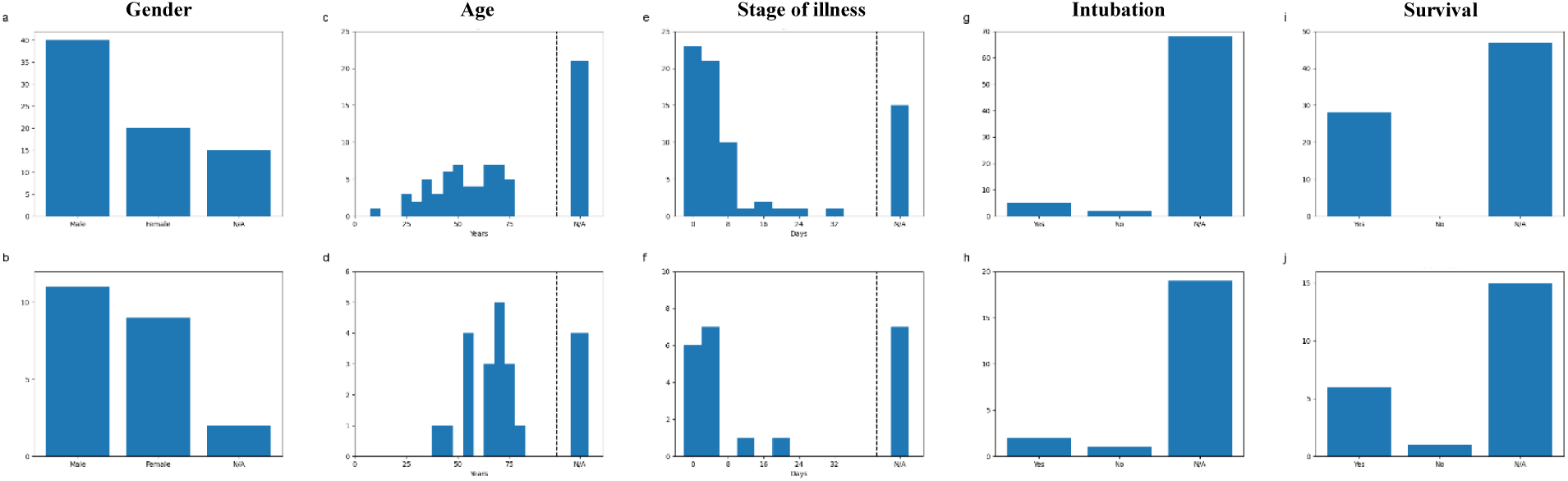
Histograms of confounding factors. Top row: L phenotype. Bottom row: H phenotype. There are more L-type than H-type patients, but their distributions are similar.

With the current data, it would be beneficial to repeat the training for more specific regions of interest and to use a wider range of metrics, e.g. structural similarity^27^ and image statistics^28^. Likewise, it would help to leverage an additional viewpoint, such as the lateral (side) x-ray images commonly taken at the same time as the frontal view. Lateral x-rays are particularly useful for assessing the air spaces behind the breastbone and heart^13^ and provide an extreme form of limited-angle tomography^29^, giving much-needed perspective for volume estimation. Finally, it is important to correlate the x-ray images with data from other observations as well^30^, such as cardiovascular^24^ and renal^31^ metrics, the improvement of many patients via prone positioning (laying on the stomach)^32^, and acoustic signatures of wheezing, gasping, and coughing^33^. Far more data and clinical studies are needed, but the classification of patients should start now.

## Methods

We used the publicly available COVIDx dataset^10^, which is a combination of COVID-19 image data collection^12^ and the RSNA pneumonia dataset^11^. To date, the training set includes posteroanterior (PA) x-ray images from 7966 pneumonia-positive patients and 8066 “normal” patients (pneumonia-negative and COVID-19-negative patients) but only 98 images of COVID-19-positive patients. The test set consists of 100 normal, 100 pneumonia, and 10 COVID-19 patients. For the t-SNE and UMAP analysis, we employed 308 normal, 308 pneumonia, and 98 COVID-19 x-ray images.

### Transformation and Preprocessing

Images are resized such that the lowest dimension contains 256 pixels and then center-cropped for feature analysis. For fine-tuning the network, we augmented the data with random jitter in brightness, random crops, and horizontal and vertical flips.

### Neural Network

We use a DenseNet-121 architecture^34^ trained on the ImageNet database^35^, performed using the PyTorch deep learning library^36^. The network uses Adam optimization^37^ with an initial learning rate of 0.0001. It is then trained for a total of 10 epochs, with the learning rate decreasing by a factor of 0.1 every 3 epochs. For evaluation, we employ a five-crop criterion in which we take four corner crops and one center crop of the image and make decisions based on majority rule. The fine-tuned network showed a sensitivity of 80% for COVID-19 detection and 93% for pneumonia detection.

### Clustering Algorithms

Image features output from the pre-trained network are grouped using the t-SNE^17^ and UMAP^19^ clustering algorithms. To confirm consistency, the t-SNE perplexity value was varied from 20 to 100 and the number of iterations was varied from 1000 to 5000; for the final results, the perplexity value is set to 50 and the number of iterations is set to 1000. For the UMAP analysis, spread values of 1–3 were considered; for the final results, the value is set to 2. To obtain a consistent topology, the UMAP distance parameter is tuned to 0.1.

## Data Availability

The data employed in the study are publicly available in the links:
1. COVID-19 Image Data Collection, https://github.com/ieee8023/covid-chestxray-dataset
2. RSNA Pneumonia Detection Challange, https://www.kaggle.com/c/rsna-pneumonia-detection-challenge

## Conflicts of Interest

The authors have no conflict of interest relevant to this article.

## Acknowledgements

The authors thank DARPA and the AFOSR for general support.

## Supplementary Information for

Distinguishing L and H phenotypes of Covid-19 using a single x-ray image

Islam, Mohammad Tariqul

Fleischer, Jason W.

Department of Electrical Engineering, Princeton University, Princeton, NJ 08540

This PDF file includes:

Supplementary Text

Supplementary Figures

Supplementary References

This Supplementary Information analyzes the data in more detail. Part 1 discusses the dimension-reduction methods of t-SNE^1^ and UMAP^2^. Part 2 describes the discovery by UMAP of a mislabeled x-ray image in the COVIDx dataset^3^. Part 3 provides a deeper discussion of existing points in the UMAP classification and Part 4 explores predictive testing by UMAP for new data points.

## 1. t-SNE and UMAP algorithms

Both t-SNE and UMAP reduce the high-dimensional dataset X to a low-dimensional space Y (typically, the target space is two-dimensional). In t-SNE, image pairs are assigned a joint probability and grouped by similarity^1^. The algorithm is non-parametric, i.e. it does not learn a mapping *f*: *X* → *Y*. Consequently^1,4^,

1. there may be no meaning to the placement of clusters;
2. there may be no meaning to the distance between clusters; and
3. the algorithm cannot simply add new data points to an existing plot; it must be re-run on the expanded set when new data points are added.

The first two properties mean that multiple runs of t-SNE may result in multiple output geometries. In our case, we observe two different manifestations, depending on the (random) initial condition chosen to start the algorithm: half the time it places a COVID-19 cluster between those of pneumonia and normal x-rays (Fig. S1a), half the time it places the COVID-19 cluster above the normal one (Fig. S1b). The third property can be overcome by parameterizing t-SNE to accommodate new data^5^.

A more modern algorithm better suited for generalization is UMAP^2^. This algorithm provides a distance metric between points (typically Riemannian) and gives a more consistent topology over a wide range of parameters and initial conditions (Figs. 1c,d). Accordingly, we use UMAP for analysis and prediction.

**Figure S1.**
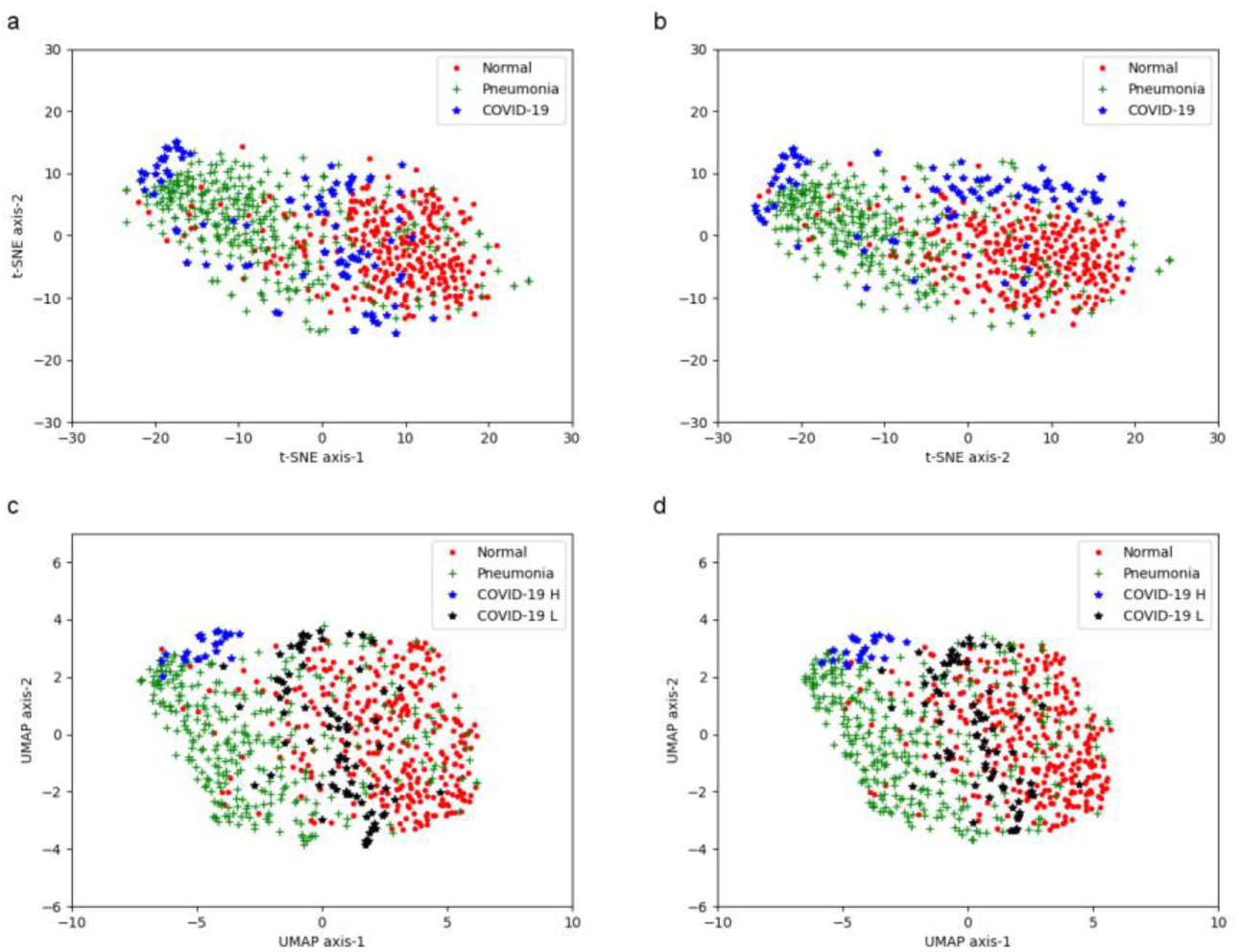
Clustering algorithms. (a,b) t-SNE clustering for two different random initializations. (a) Fig. 2c from the main text. (b) Second configuration. (c,d) UMAP for two different random initializations. In all cases, the UMAP topology remained the same.

## 2. Mislabeled data discovered by UMAP

In the original UMAP mapping, there was an extra point labeled H that appeared in the ellipse for the L phenotype. This point, shown in red in Fig. S2a, later revealed itself as an error in the COVIDx dataset^3,6^: it was labeled as a frontal (PA) chest x-ray in the physician notes, when in fact it is a lateral (side) x-ray image (Fig. S2b). We removed this data point in subsequent analysis.

**Figure S2.**
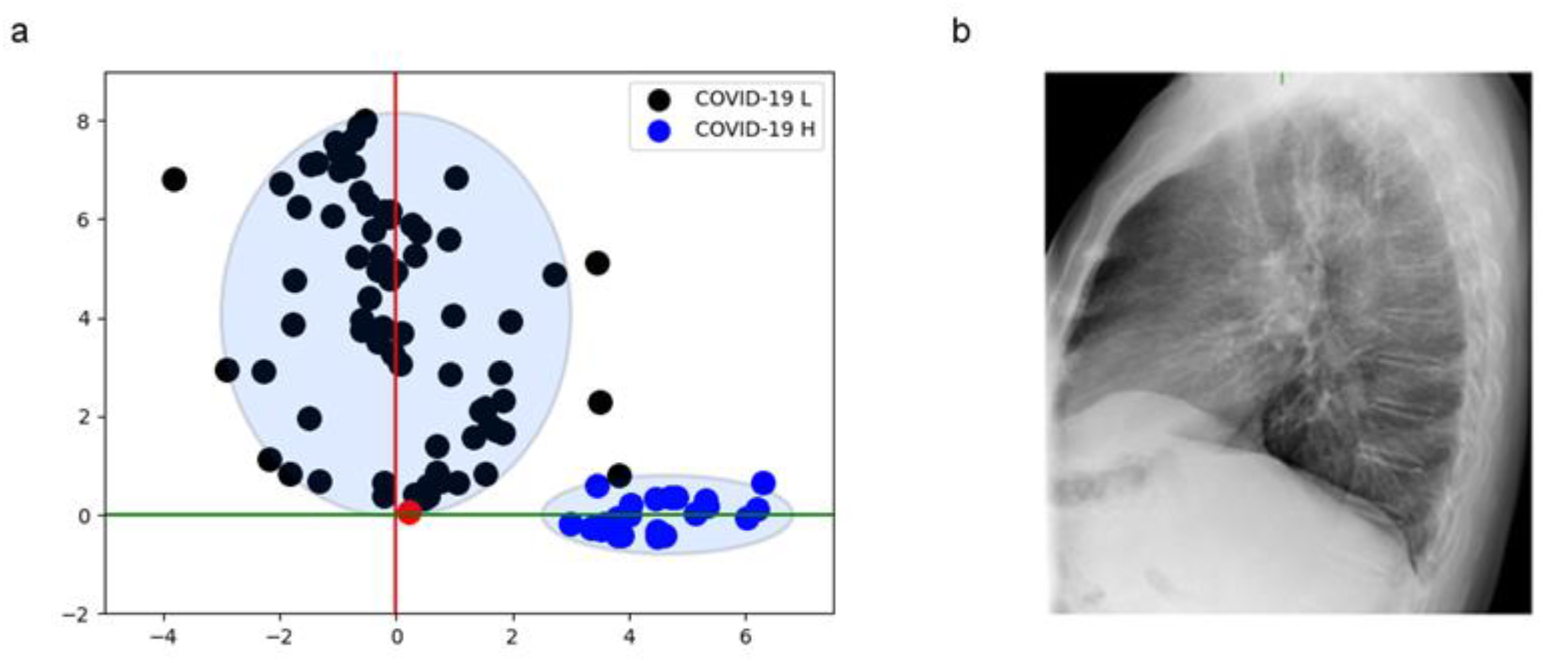
Discovery of mislabeled data. (a) Fig. 3a from main text with mislabeled data point added in red. (b) Chest x-ray of mislabeled point, showing side view rather than frontal (PA) view.

## 3. Examples and Outliers From UMAP

We present some representative samples from the UMAP plot given in the main text (Fig. 3a). Shown are points within each cluster, at the boundaries, and in the outlying regions. Results for the L phenotype are given first, followed by those for the H phenotype. Under each x-ray image, we also give the available radiologist and physician notes^6^.

**Figure S3.**
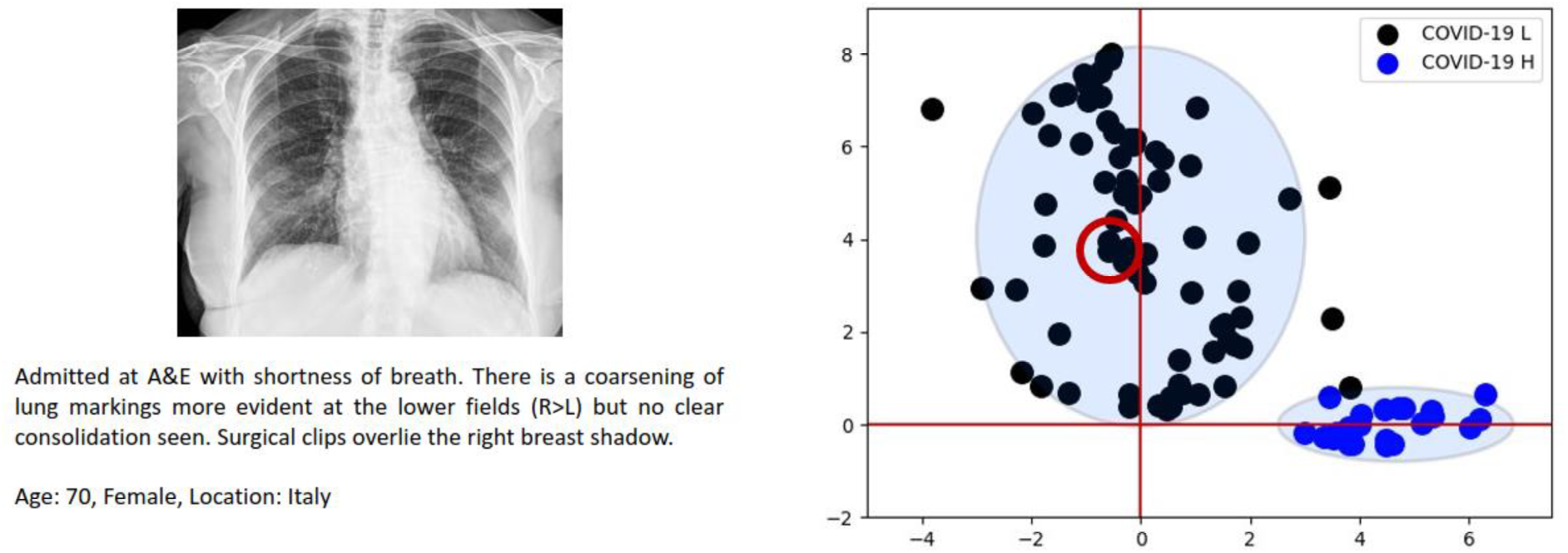
Typical x-ray image within the L cluster.

**Figure S4.**
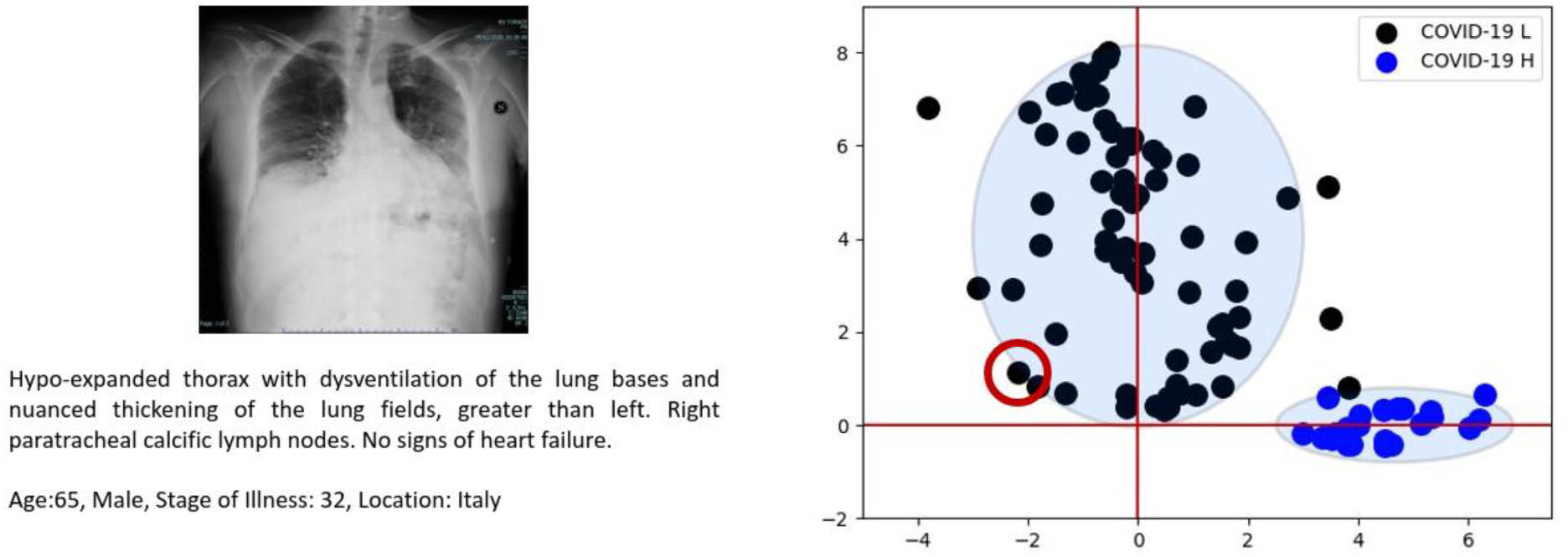
x-ray image at the boundary of the L ellipse.

**Figure S5.**
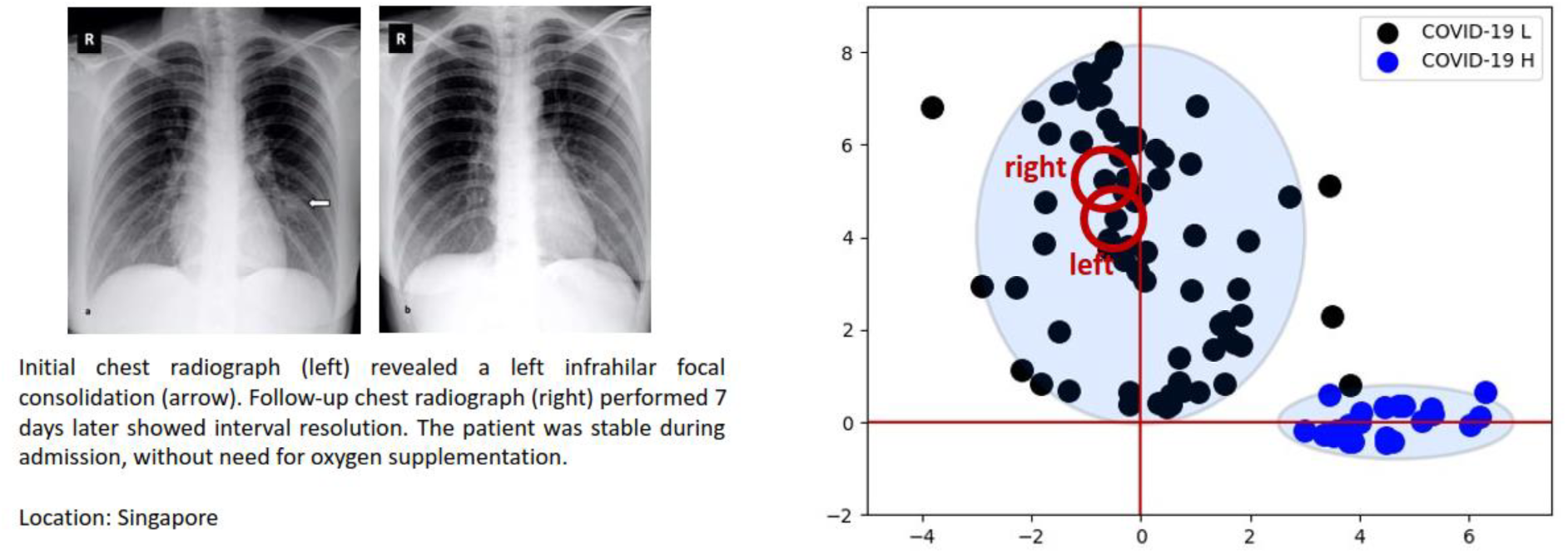
x-ray images of a stable patient in the L cluster.

**Figure S6.**
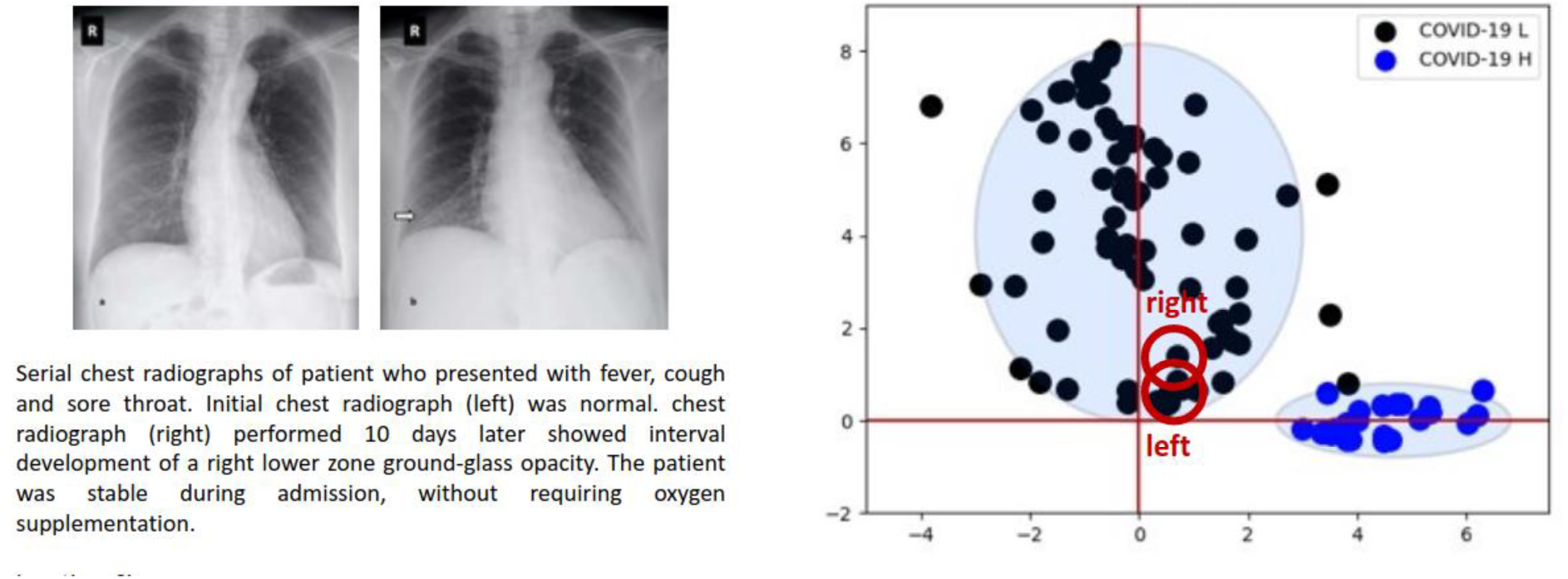
x-ray images of a stable patient in the L cluster.

**Figure S7.**
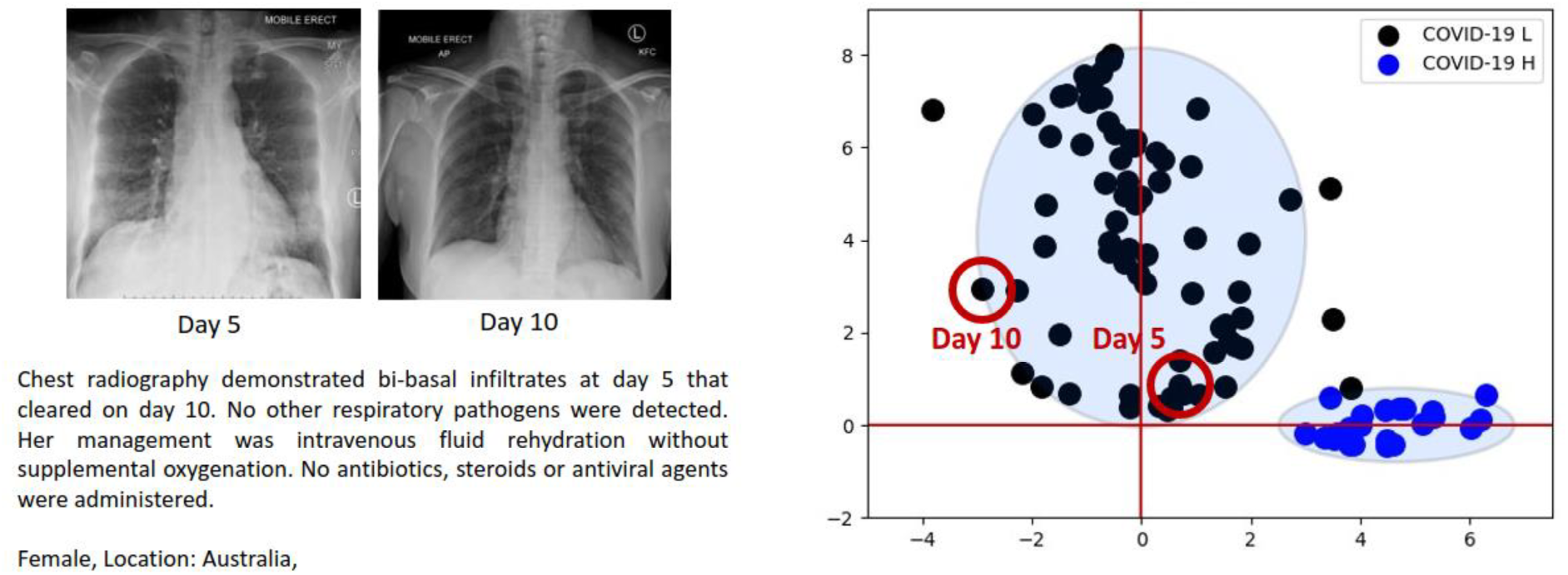
Progression of recovery in the L cluster. Points move from right to left in parameter space.

**Figure S8.**
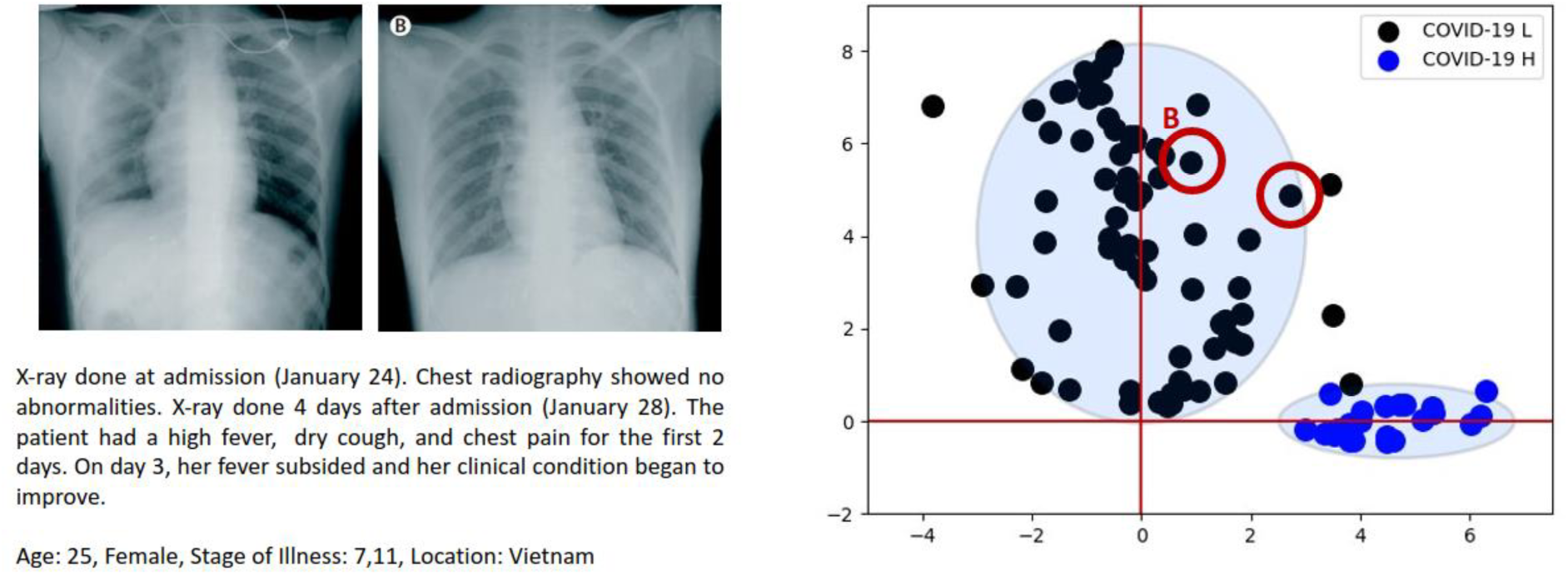
Progression of recovery in the L cluster. Points move from right to left in parameter space.

**Figure S9.**
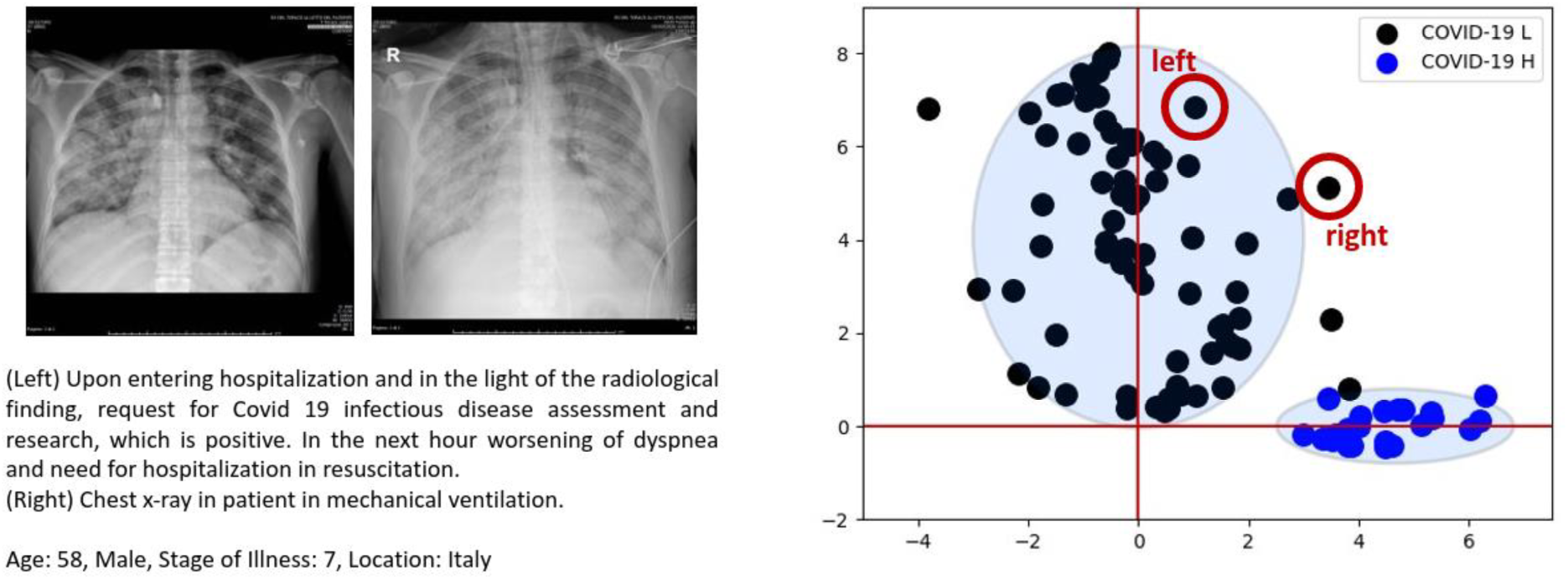
Progression of illness in the L cluster. Points move from left to right in parameter space.

**Figure S10.**
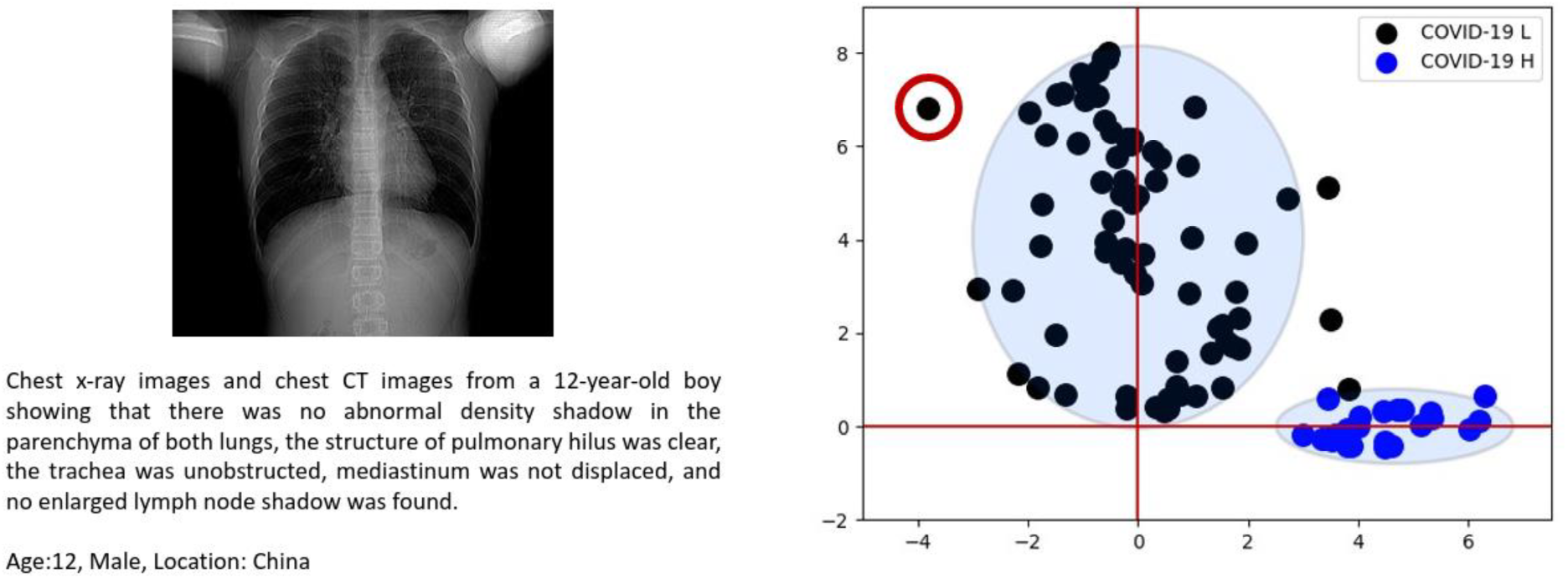
UMAP placement of the youngest COVID-19 patient in the COVIDx dataset.

**Figure S11:**
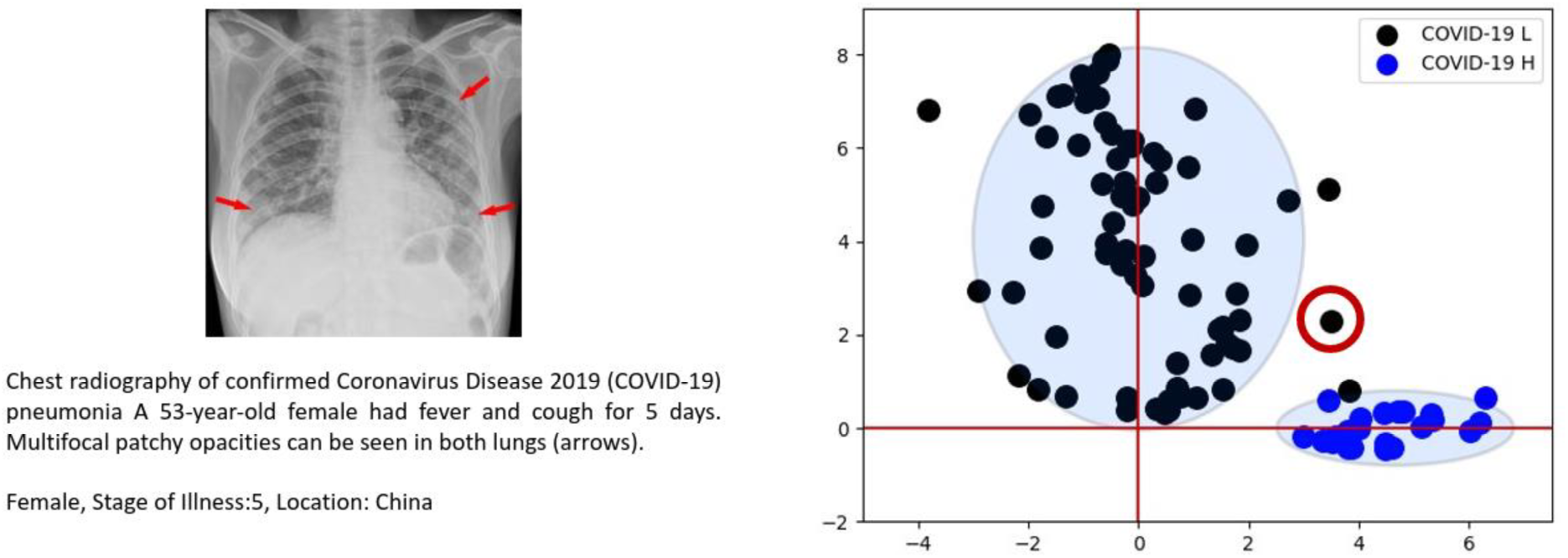
Outlier between the L and H ellipses.

**S12.**
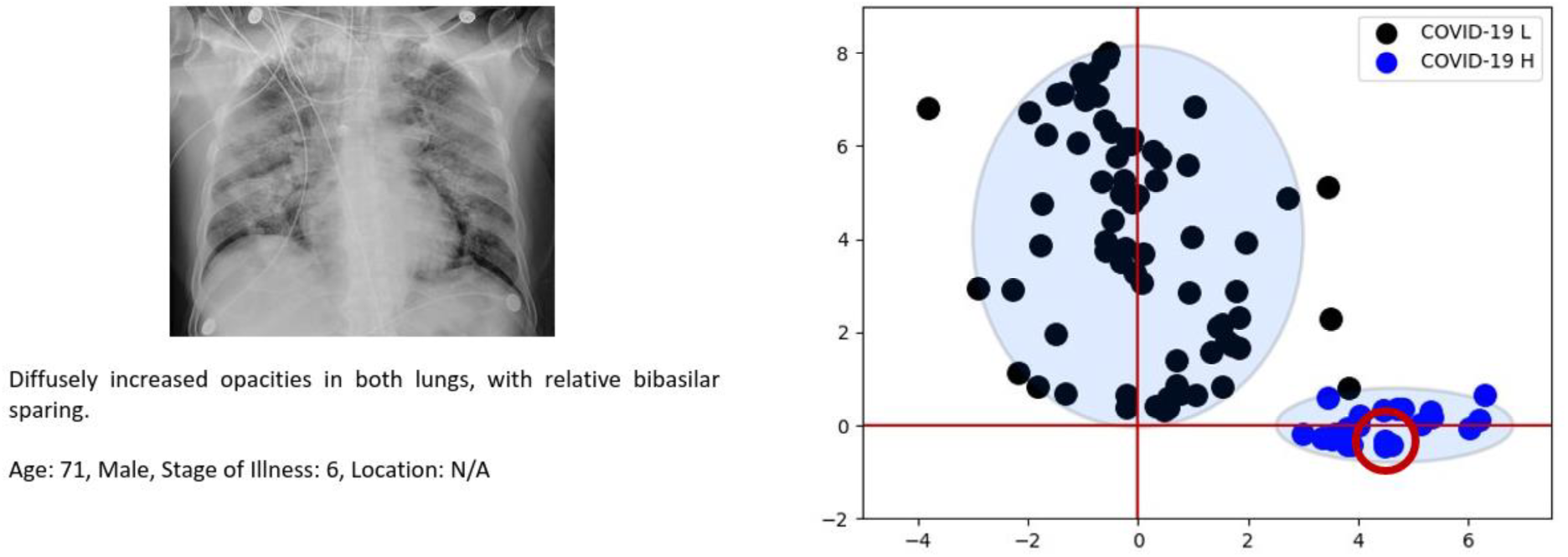
Typical x-ray image within the H cluster.

**Figure S13:**
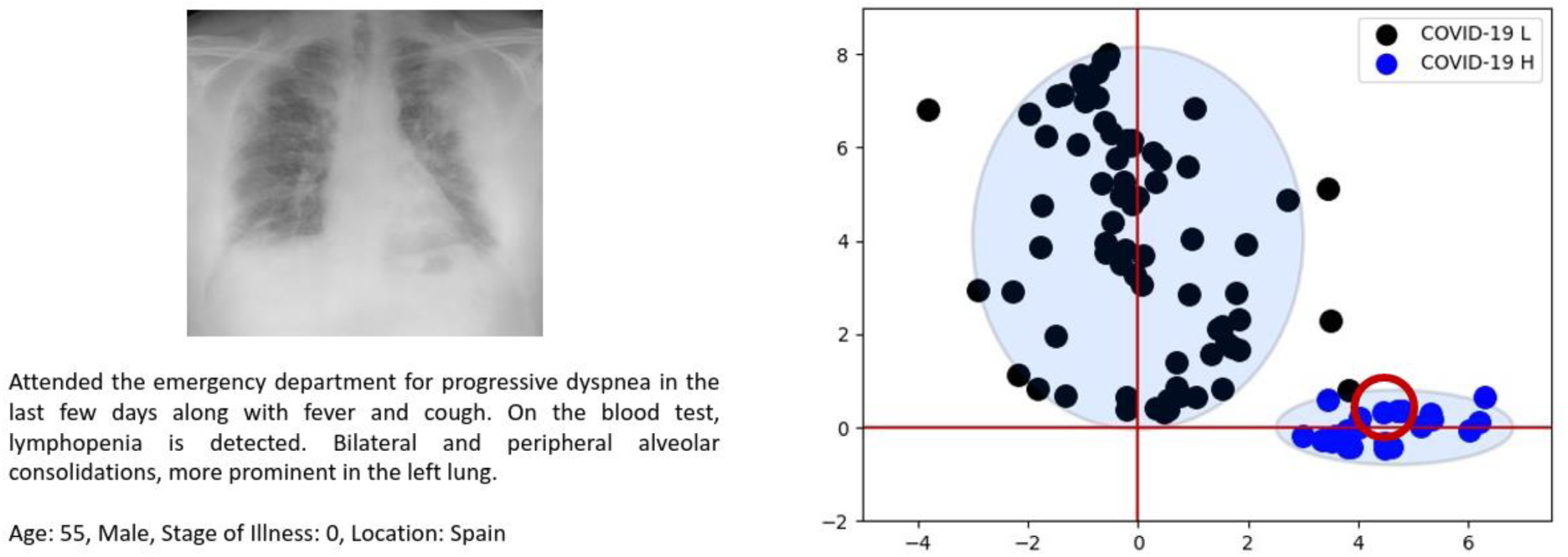
Typical x-ray image within the H cluster.

**Figure S14:**
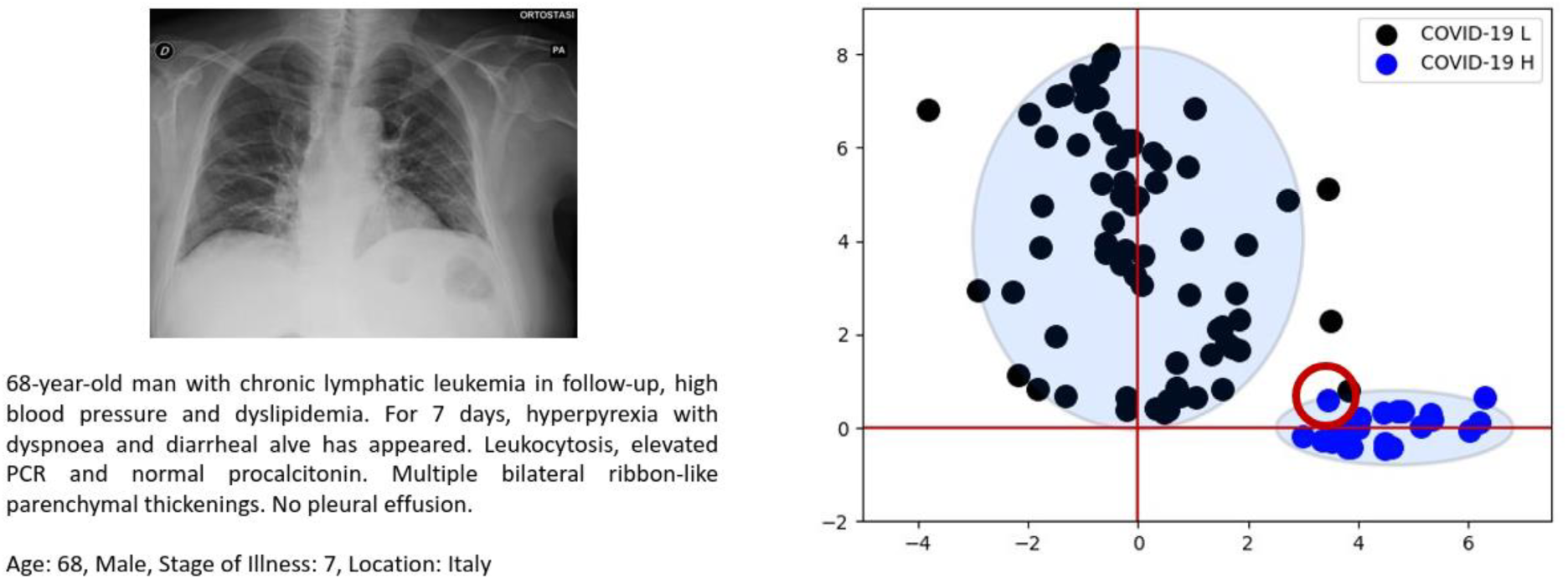
x-ray image on the boundary of the H cluster. The patient has lymphatic leukemia, which would put him in the high-risk H category regardless of imaging.

**Figure S15:**
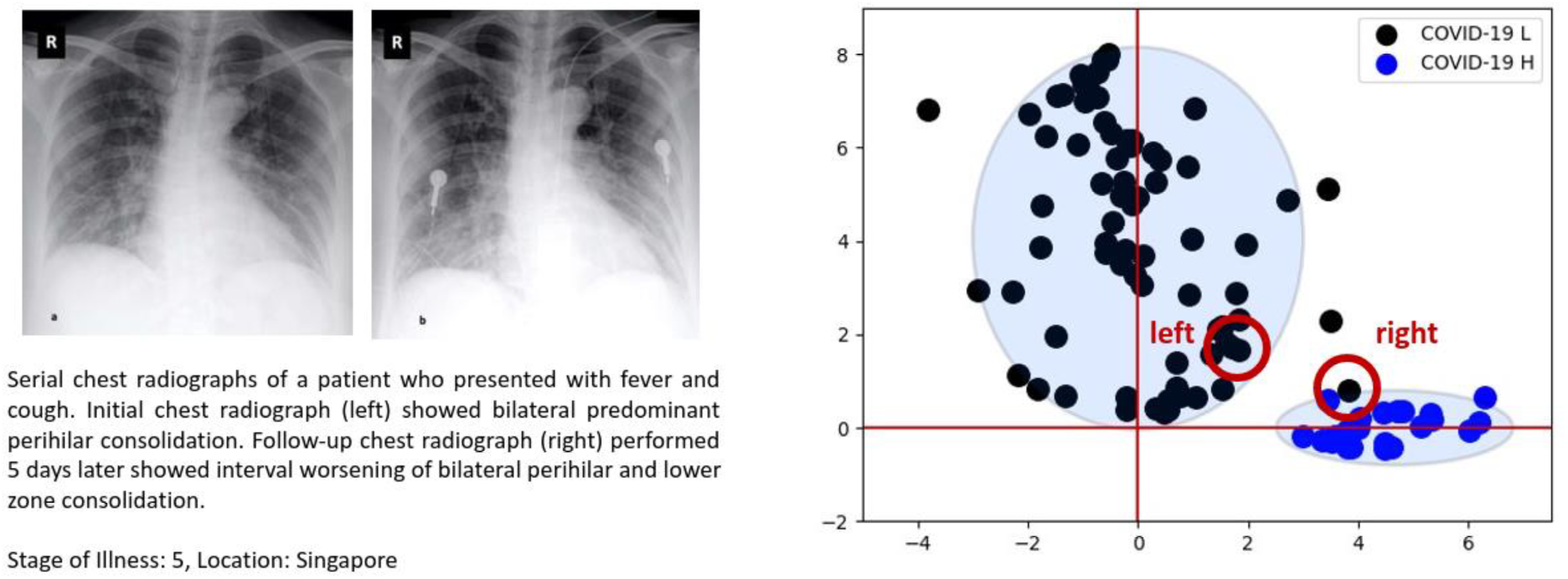
Mislabeled x-ray image in the H cluster, near the point in Fig. S14. Label from t-SNE, placement from UMAP. As before, disease worsening progresses from left to right in parameter space.

**Figure S16:**
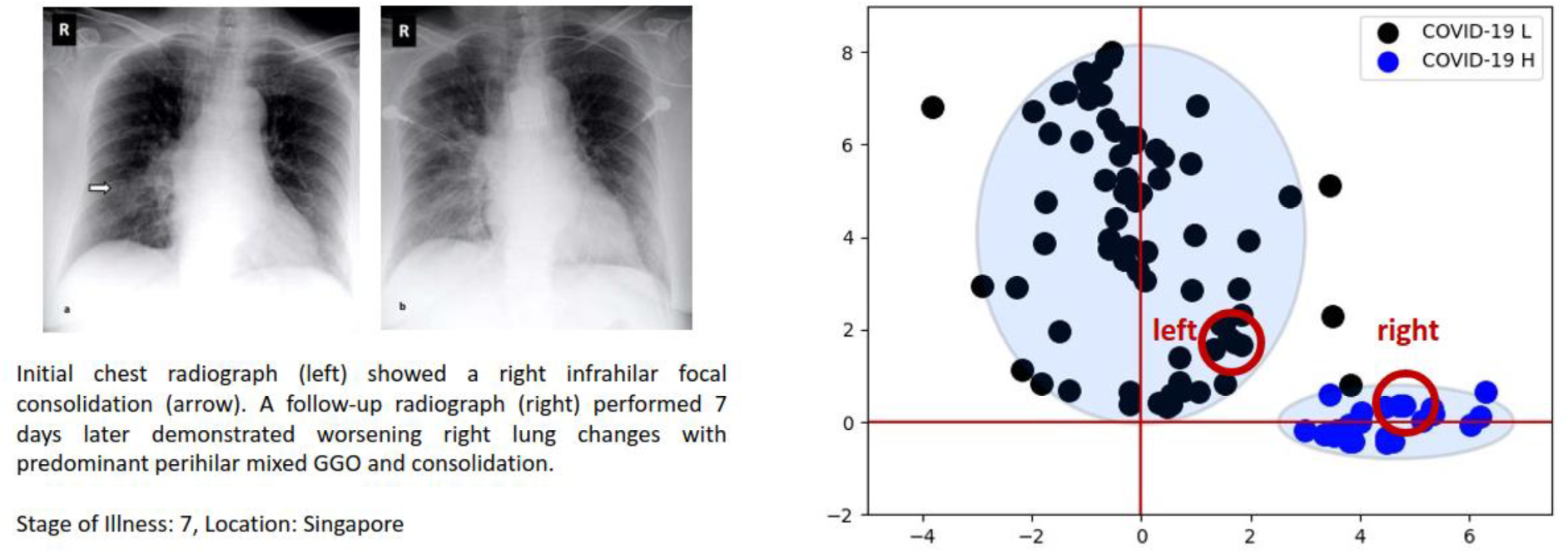
Disease progression from L cluster to H cluster.

**Figure S17:**
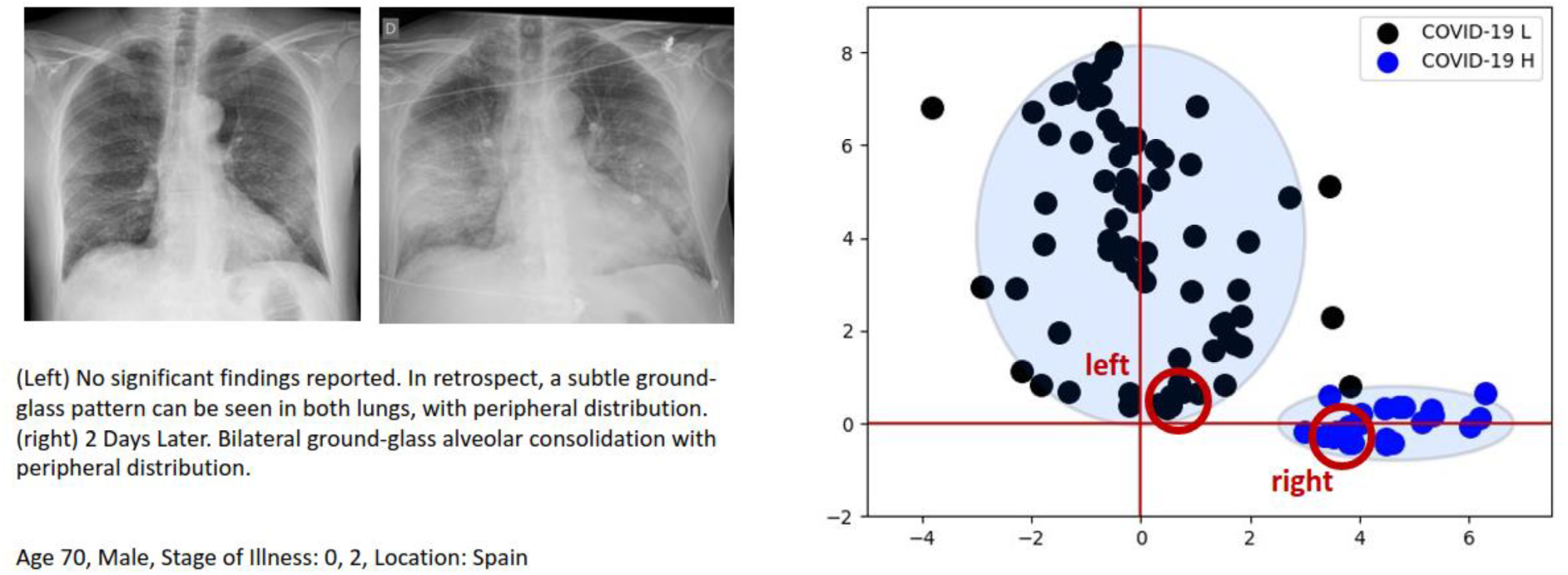
Disease progression from L cluster to H cluster.

## 4. Predictive testing of UMAP

Covid-19 x-ray data is very limited and does not allow for significant predictive testing of the network. Nevertheless, we saved a small number of x-rays from the COVIDx dataset^6^ to analyze performance on new patients, viz. 100 normal x-ray images, 100 pneumonia x-ray images, and 10 COVID-19 x-ray images. Results are shown in Figs. S18–22. As seen in previous applications of UMAP for phenotyping^7^, the new data points appear on the periphery of the previously learned mapping. As before, some patients have multiple chest x-rays, allowing comparison of different stages of illness.

**Figure S18.**
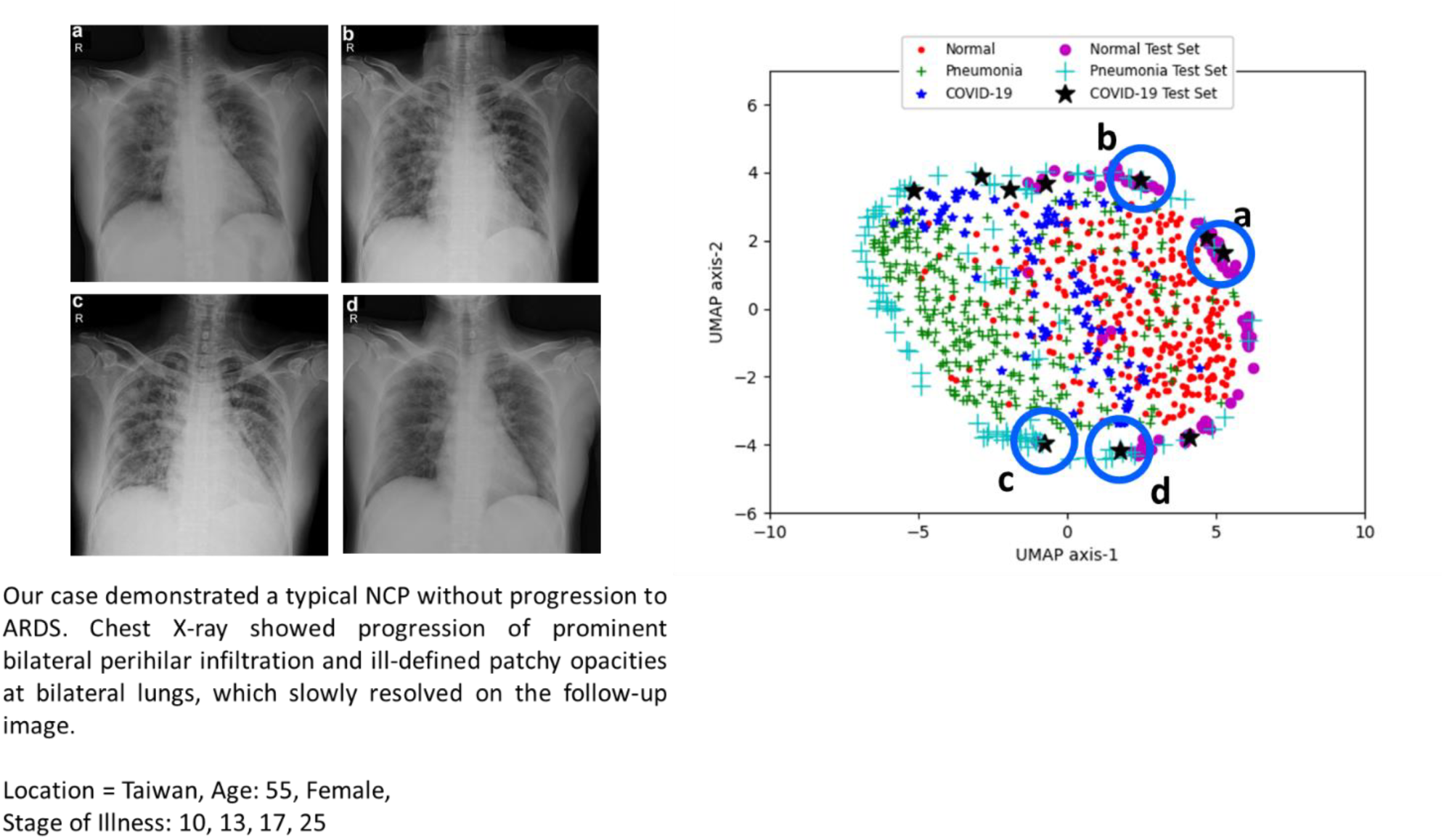
Progression of illness and recovery for L-type patient. Shown are x-rays from the same patient taken at (a) 10, (b) 13, (c) 17, and (d) 25 days after onset of illness^8^. Note that (c) corresponds to the worst stage of the disease and maps to the pneumonia-like cluster. (a,b) Worsening of disease. (c,d) Patient improvement.

**Figure S19.**
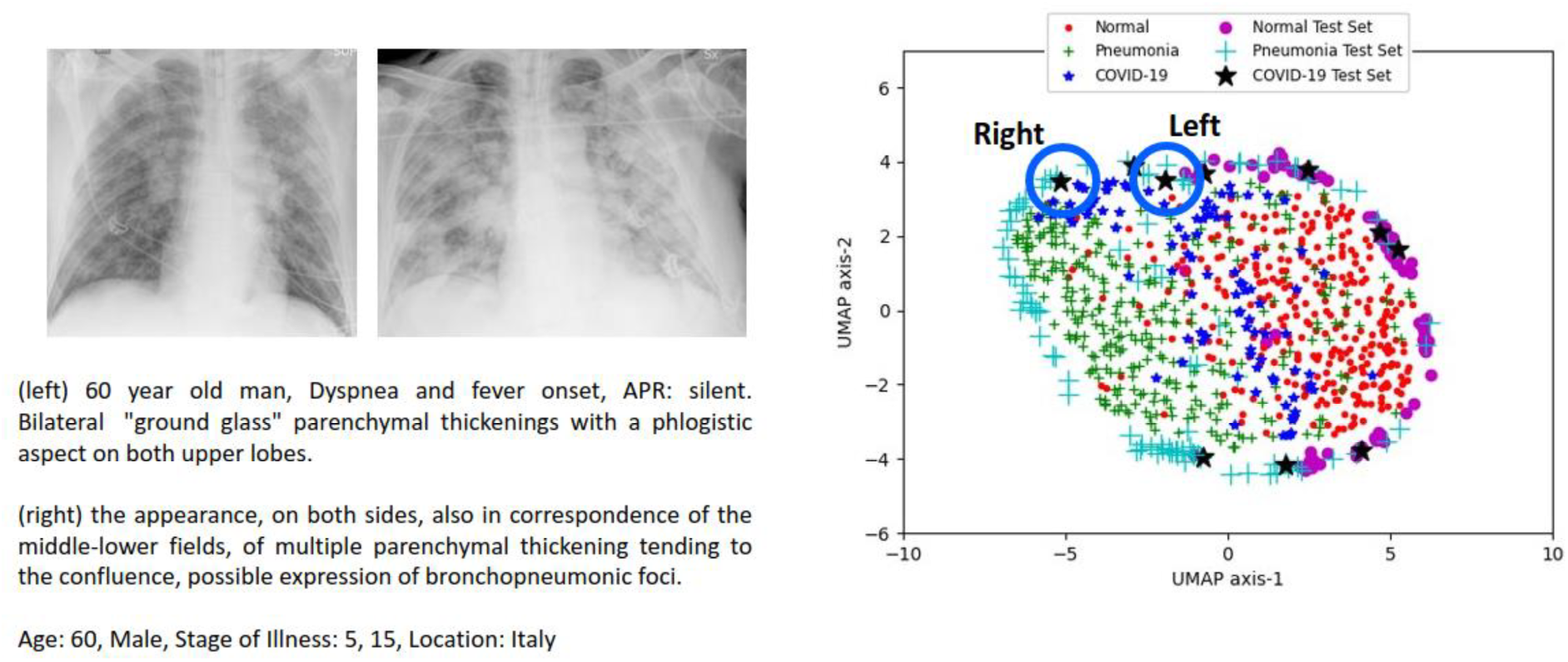
Progression of illness for H-type patient. As before, the x-ray becomes more pneumonia-like as the condition worsens.

**Figure S20.**
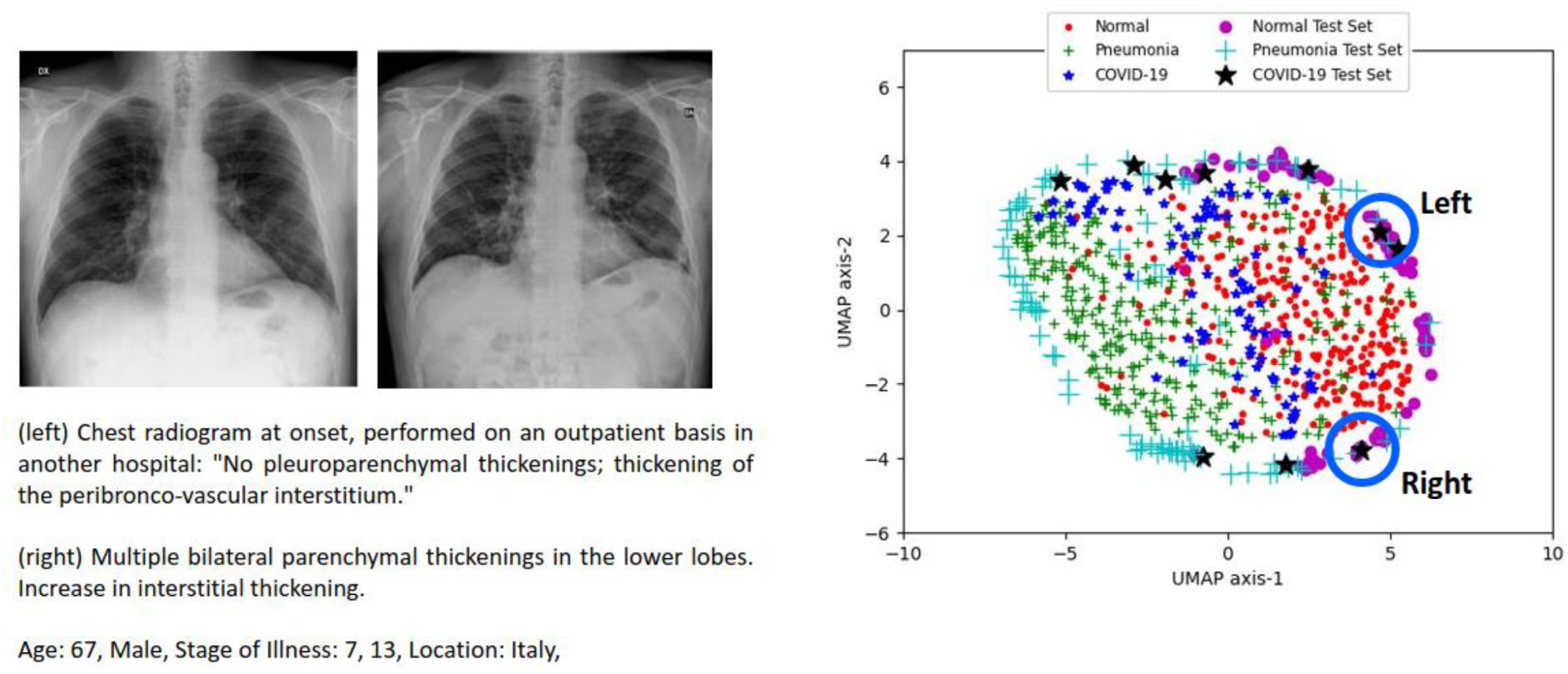
Slow progression of L-type patient.

**Figure S21.**
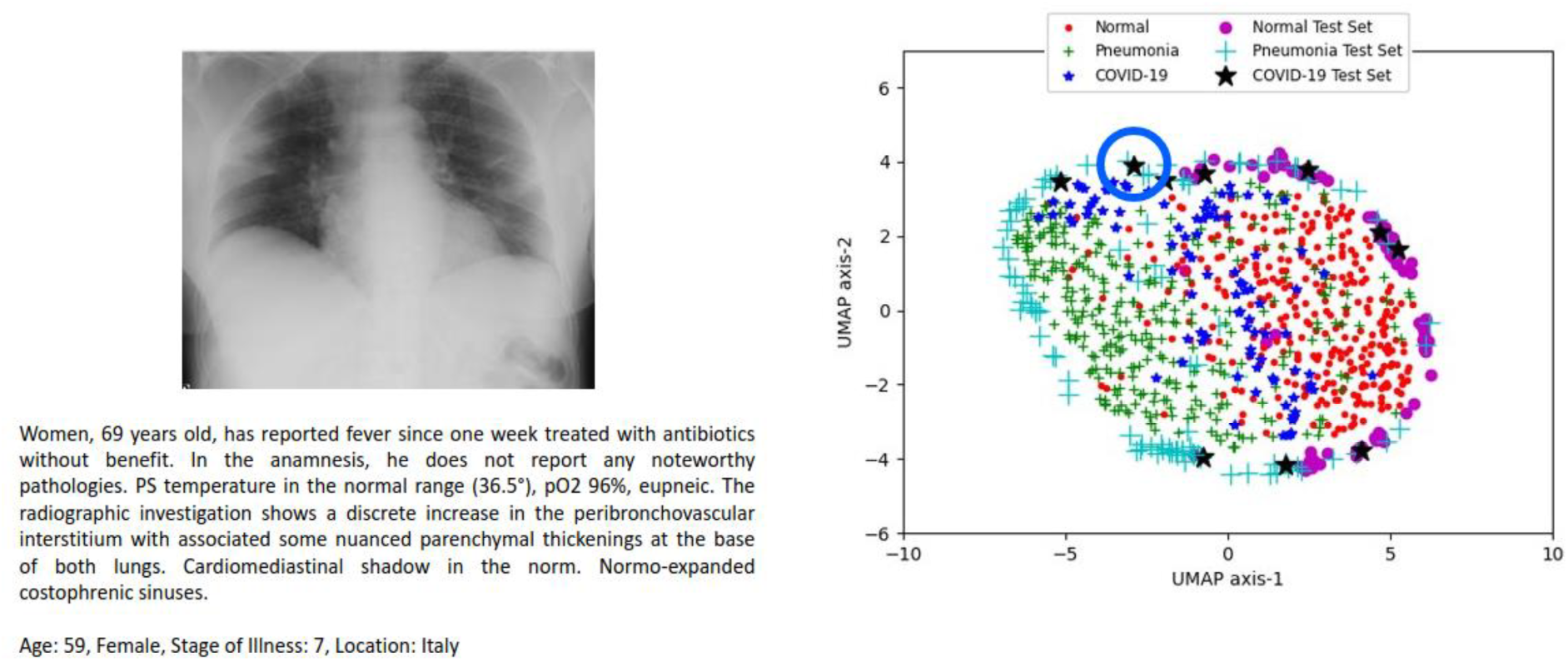
New x-ray image placed near origin of L-H axes.

**Figure S22.**
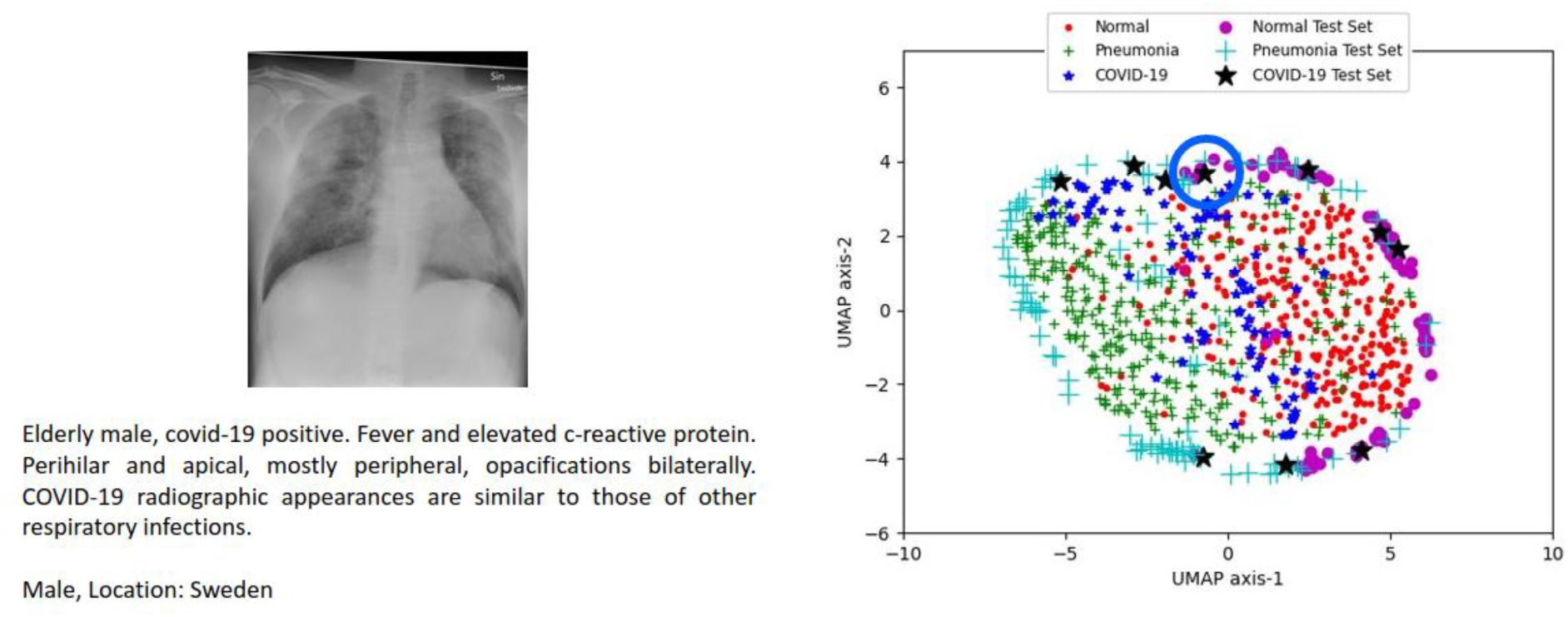
New x-ray image placed near origin of L-H axes.

## Notes

### Competing Interest Statement

The authors have declared no competing interest.

